# Economic analysis of a school-based menstrual health intervention (MENISCUS) among female adolescents in Uganda

**DOI:** 10.64898/2026.03.11.26348207

**Authors:** Stephen Lagony, Daria Bucci, Rebecca A.K. Dwommoh, Levicatus Mugenyi, Kate A Nelson, Edward Obicho, Fred Matovu, Shamirah Nakalema, Helen A. Weiss, Giulia Greco

## Abstract

Poor menstrual health (MH) has been associated with reduced participation in school activities and diminished psychosocial wellbeing among adolescent girls. Despite increasing recognition of the importance of MH interventions, there is limited economic evidence to inform large-scale adoption and financial planning.

We conducted an incremental costing analysis of an MH intervention (MENISCUS) alongside a cluster-randomized trial in 60 secondary schools in Uganda. MENISCUS delivered puberty education, a drama skit, an MH kit, pain management strategies and improvements to water, sanitation and hygiene (WASH) facilities. We categorized the provider costs into start-up and implementation, and calculated unit costs per school, per student (male and female) and per female student respectively. We modelled two potential national scale-up scenarios (basic and enhanced) to 2,995 secondary schools using government delivery structures.

The total cost of the basic scenario is US$10,224,685 and the enhanced scenario is US$16,549,123. The unit cost of scaling the intervention nationwide was estimated at US$28 per student and US$58 per female student (basic scenario) and US$46 per student and US$95 per female student (enhanced scenario). The primary cost drivers were the MH kit and associated training, followed by pain management activities and improvements to WASH facilities. The enhanced scenario generated a higher unit cost per student and unit cost per female student due to additional components. Compared with trial costs, unit costs were lower in national scale-up, demonstrating economies of scale.

This study provides the first economic analysis of a potential national implementation of a school-based MH intervention in a low-resource setting. The findings provide critical benchmarks for governments seeking to integrate MH into national education curriculum and inform future investment decisions in adolescent health.

## Introduction

Poor menstrual health (MH) is a major global health concern with adverse outcomes for quality of life (1–3). Studies in low-and middle-income countries (LMICs) show that poor MH can be associated with school absenteeism, low engagement and decreased participation in activities among in-school female adolescents (4–9). School-based MH interventions have improved adolescents’ MH knowledge, attitudes and practices (10), but there is less evidence on the impact on improving psychosocial outcomes, mental health and well-being (11–16).

Costing and economic analysis studies are needed to inform policy, resource planning, and program implementation of MH interventions. These studies identify key cost drivers and support priority setting by enabling policy makers to assess affordability and budgetary feasibility within constrained health and education systems (17,18). However, few studies have evaluated the cost of MH interventions, including for scale-up and financial sustainability (19). There are also few studies on the longer-term economic value of MH programs on improving health, education, and psychosocial outcomes. A study in Kenya conducted a cost-benefit and cost-effectiveness analysis of providing menstrual pads and menstrual cups to schoolgirls in rural Kenya (20). The annual cost of providing menstrual cups and menstrual pads was US$3.27 and US$24.00 respectively per direct recipient. The authors concluded that the menstrual cup is likely to be a cost-effective solution for MH in low-resource settings, with one menstrual cup predicted to last 10 years (20). In 2022-2023, we conducted a cluster-randomized controlled trial of a multi-component intervention (MENISCUS) in 60 Ugandan secondary schools (16,21). In the 30 intervention schools, we estimated the total cost of setting up the intervention at US$40,990 and the annual implementation cost at US$181,503. The annual economic cost was US$44 per student and US$85 per female student (16). To our knowledge, these are the only two studies that have conducted costing analyses and an economic evaluation of a school-based MH intervention in an LMIC.

In this paper, we model two potential national scale-up scenarios based on the MENISCUS intervention. We use data from the completed trial that evaluated the impact of the intervention on educational, quality of life and mental health outcomes, as well as menstrual health outcomes. (16,21). Specifically, we ask, what would it cost to scale up the MENISCUS intervention to national level under two different implementation scenarios? This information is necessary for policy makers because it provides real world estimates of financial implications under real world delivery systems. We did not conduct a cost-effectiveness analysis as part of the MENISCUS trial due to the lack of evidence of an intervention effect on the primary educational mental health outcomes (21). However, there was strong evidence of an intervention effect on menstrual health secondary outcomes, of interest to policy makers.

## Methods

### Study Design

The trial design and outcome evaluation have been published elsewhere (16,21). In brief, 60 secondary schools were randomized 1:1 to receive either the MENISCUS intervention package or optimized usual care. Schools were eligible if they were in Wakiso or Kalungu districts, mixed-gender, had both day and boarding students, and basic water, sanitation and hygiene (WASH) facilities. Baseline assessments were conducted from 21^st^ March 2022 to 5^th^ July 2022, and endline assessments from 5^th^ June 2023 to 22^nd^ August 2023. We recruited study participants during baseline and sought written informed assent from female students younger than 18 years, a long with parental consent. We also sought electronic written informed consent for female students aged 18 years or older. We obtained written school-level consent from the headteacher or a representative in 60 schools randomly sampled from those eligible.

### Study Population

The MENISCUS study targeted female and male students enrolled in secondary schools in Kalungu and Wakiso with a primary focus on Senior 2 (S2). A total of 3,841 female students and 874 male students were enrolled in the study. The median age of female participants was 15 –16 years (iQR 15 –16), while male participants had a median age of 16 years (IQR 15 –17). Approximately 55% of study participants were day students and 45% boarding students. Religious affiliation of the participants was largely diverse, with roughly 30% identifying as Catholics, 37-40% as Protestants/Born Again/Seventh Day Adventist, and approximately 27-31% as Muslim. Approximately 69% of female participants identified as Muganda, which reflects the dominant ethnic tribe in the region. Household sizes were generally large with roughly 40% of female participants reporting households of eight or more members. Socio economic status, derived from an asset list, was evenly distributed across lowest, medium and highest positions (approximately one third in each category) (16).

### The MENISCUS intervention

The intervention development was guided by Social Cognitive Theory (22) and was implemented in collaboration with WoMena Uganda, a local non-government organization (NGO). The intervention package has been described in detail elsewhere (21), and included i) puberty education, ii) implementation of a drama skit, iii) a MH kit that contained 5 re-usable Ugandan-manufactured pads, underwear, a towel, water bottle, a bar of soap and an option of a menstrual cup with a storage container, iv) pain management and v) basic improvements to school WASH facilities. The components were delivered to both male and female students in one school year (S2), except for the MH kit, analgesics and pain relief training, which were offered only to female students. Female students had the option to receive a medical-grade silicone re-usable menstrual cup with a container for disinfection and storage. An MH action group was set up in each school to coordinate and maintain the intervention. The group comprised 6–8 teachers, students and parents selected by school management. Optimized usual care included the provision of government guidelines on MH management, provision of the National Sexuality Education Framework (23) and the provision of a copy of the government menstrual management reader to all male and female S2 students in trial schools.

The participants in the intervention group reported fewer unmet menstrual needs, greater self-efficacy to manage menstruation, improved use of effective pain management, greater knowledge of puberty and menstruation and improved positive attitudes towards menstruation (16).

### Collection of cost data for the MENISCUS trial

We collected cost data in June 2023 using a combination of bottom-up and top-down micro costing approaches. Data sources included financial records, project accounts, monitoring and evaluation (M&E) registers, and staff timesheets from WoMena Uganda (main implementing partner) and the MRC/UVRI and LSHTM Uganda Research Unit (research partner).

Costs were incurred from January 2021 to June 2023 and were categorized into start-up and implementation phases. The start-up period (January 2021 to May 2022) covered planning meetings, exploratory meetings, stakeholder meetings, training of internal trainers, development of Training of Trainers (ToTs) handbook and procurement of equipment. The implementation phase started after randomization from June 2022 to June 2023. Costs were annuitized over their expected useful lives: 18 months for start-up activities and 1 year for implementation activities. For policy relevance, analyses were conducted from a providers’ perspective and included both financial costs (actual expenditures) and economic costs (opportunity costs of all resources used). All research costs were excluded.

### Cost Components for the MENSICUS trial

We estimated full-time equivalents (FTEs) for all WoMena staff involved in implementation of the intervention based on timesheet data and interviews with staff. Positions included project manager, Monitoring & Evaluation personnel, project officers, finance and administrative personnel. We allocated time spent on core versus support activities using a top-down allocation factor based on the number of activities conducted during each phase.

Capital costs such as furniture and equipment were obtained from WoMena’s asset registry, annualized, and adjusted for inflation. Rental costs for office space were allocated based on the proportion of surface area used by the intervention: two out of five rooms were fully allocated to MENISCUS activities. We assumed that the space was used daily and fully dedicated to intervention tasks. Overhead costs were allocated using a top-down micro-costing approach. These included building maintenance and utilities, which were proportionally distributed based on the surface area occupied by the intervention.

Recurrent costs included venue hire for different MENISCUS trainings, meals and refreshments, printing training handbooks, office and training supplies and menstrual health kits. Charges for mobile money transactions used during implementation were also included. These charges were fees levied for withdrawing money using mobile money, a popular digital financial service that facilitates financial transactions using a mobile phone. Supplies were directly allocated to intervention or administrative functions based on actual usage. Supplies included procurement and distribution of materials such as MH kits, WASH infrastructure, implementation of a drama skit, puberty education training and provision of analgesics. Transport costs (e.g., fuel, car hire, transport reimbursements) and accommodation expenses were extracted from financial records and allocated to specific activities based on M&E documentation and activity logs (16).

### Costing Approach

We estimated the incremental total costs associated with the set-up and implementation of the MENSICUS intervention in the trial setting and modelled two potential national scale-up scenarios. The costs of providing optimized usual care were excluded from the intervention costs to estimate the incremental costs attributed to the delivery of the MENISCUS intervention.

We present scale-up costs using two scenarios: (1) Basic scale up scenario and (2) Enhanced scale up scenario. These scenarios use the Government of Uganda model of delivery where the intervention will be delivered using existing government systems, structures and personnel as opposed to a non-government approach. We report start-up costs and implementation costs for the trial in 2023 US dollars.

We followed the Reference Case approach for costing, recommended by the International Decision Support Initiative (iDSI) and the Consolidated Health Economic Evaluation Reporting Standards (CHEERS) (24,25). We used a micro-costing methodology (26–28), consistent with best practices for cost analysis of health and social interventions in LMICs, drawing on the Guidelines for Conducting Cost Analyses of Interventions to Prevent Violence Against Women and Girls (VAWG) (29), which align with both the iDSI (24) and the Global Health Costing Consortium (GHCC) Reference Case (30). These guidelines include openly available Excel-based costing tools, which we adapted for our intervention.

### Unit Costs

Unit costs were calculated by dividing total (fixed and variable) costs by three output measures: (1) number of S2 students (male and female) (N=4,120), (2) number of S2 female students (N=2,138), and (3) number of schools implementing the intervention (N=30). Costs were recorded in Ugandan Shillings (UGX) and converted to USD using the Bank of Uganda exchange rate for the midpoint of each phase: UGX 3,724.39/USD (30 May 2022) for the start-up phase and UGX 3,662.13/USD (30 June 2023) for the implementation phase. All costs were adjusted to 2023 price levels using the GDP deflator from the World Bank (31,32). Discounting was applied using phase-specific rates: 7% for the start-up period (January 2021–May 2022) and 11% for the implementation period (June 2022–June 2023), based on Bank of Uganda interest policy rates as recommended by World Health Organization (WHO) for developing countries (16,33).

### Potential national scale-up scenarios

Based on the costing analysis of the MENISCUS intervention within the trial setting, we modelled two scenarios (basic and enhanced) in Microsoft Excel for potential national scale-up using the Government of Uganda model of delivery. These scenarios assume the MENISCUS intervention will be delivered across all the 2,995 secondary schools in 146 districts of Uganda. We modified the intervention package for each of the scenarios using an elicitation process involving five core members of the MENISCUS team who led the design and implementation of the MENISCUS trial. The team employed a subjective scoring framework using two criteria: centrality to the theory of change and acceptability of the intervention items based on qualitative interviews conducted at endline during the implementation of the MENISCUS trial (21). The MENISCUS intervention items were scored across the two criteria from 1 to 4 where 1 denotes the most favorable and 4 the least preferred option. Scenario 1 (the basic scale up scenario) comprises items/activities that scored 1 for both criteria and Scenario 2 (the enhanced scale up scenario) additional included items/activities that scored 1 and 2, or 2 and 2 and 2 (Table 1). The basic scenario includes puberty education (training teachers on how to deliver the Ministry of Education and Sports’ (MoES) training on menstrual health management), MH kit and related training (training of trainers (ToTs) and training of beneficiaries), training on pain management techniques, WASH improvements (fixing of broken toilet doors, installation of toilet door locks, installation of toilet paper cages and procurement of WASH stands, drums and basins) and implementation of the drama skit (printing the drama script and drama skit rehearsals). The enhanced scale up scenario includes all elements of the basic scenario described above as well as the MH action group (training of MH action group members), training of female care givers, provision of five reusable menstrual pads (Afripads), provision of underwear for female students and provision of six tablets of ibuprofen per female student per month (Table 1).

**Table 1:** Subjective scores for MENISCUS items and activities, by acceptability of the intervention and centrality to the theory of change, to inform the composition of the two potential MENISCUS scale up scenarios.

|  | **MENISCUS Items and Activities** | **Acceptability**<br>**of the**<br>**intervention** | **Centrality to**<br>**the theory of**<br>**change** | **Basic**<br>**scenario** | **Enhanced**<br>**Scenario** |
| --- | --- | --- | --- | --- | --- |
| **1** | **Puberty Education** |  |  |  |  |
| a) | Training teachers to deliver the Ministry of Education and Sports' (MoES) training on MH management | 1 | 1 | ✓ | ✓ |
| **2** | **MH Action Group** |  |  |  |  |
| a) | Training of MH action group members | 2 | 1 |  | ✓ |
| **3** | **MH kit & training** |  |  |  |  |
| a) | Training of trainers (ToTs) | 1 | 1 | ✓ | ✓ |
| b) | Training of Beneficiaries (ToBs) | 1 | 1 | ✓ | ✓ |
| c) | Training of female care givers | 2 | 2 |  | ✓ |
| d) | Afripads – reusable pads | 2 | 2 |  | ✓ |
| e) | Afripads – Underwear for female students | 2 | 2 |  | ✓ |
| f) | MH cup (ruby cup) | 3 | 3 | - | - |
| g) | Metallic container for the cup | 4 | 4 | - | - |
| h) | A bar of laundry soap | 4 | 4 | - | - |
| i) | Towel | 4 | 4 | - | - |
| j) | Drawback string bag | 4 | 4 | - | - |
| **4** | **Pain Management** |  |  |  |  |
| a) | Training on pain management | 1 | 1 | ✓ | ✓ |
| b) | Provision of ibuprofen | 2 | 2 |  | ✓ |
| c) | Provision of paracetamol | 4 | 4 | - | - |
| **5** | **WASH improvements** |  |  |  |  |
| a) | Fixing of broken toilet doors | 1 | 1 | ✓ | ✓ |
| b) | Installation of toilet door locks | 1 | 1 | ✓ | ✓ |
| c) | Installation of toilet paper cages | 1 | 1 | ✓ | ✓ |
| d) | Procurement of WASH stands, drums and basins | 1 | 1 | ✓ | ✓ |
| e) | Sanitary bins | 3 | 2 | - | - |
| f) | Provision of hand washing soap | 3 | 3 | - | - |
| **6** | **Drama skit** |  |  |  |  |
| a) | Printing the drama script | 1 | 1 | ✓ | ✓ |
| b) | Drama skit rehearsal | 1 | 1 | ✓ | ✓ |
| c) | Drama skit performance | 2 | 1 | ✓ | ✓ |

The target population for scale-up was defined as Senior 1 (S1) students, based on enrollment figures and the number of secondary schools nationally, as reported by Ministry of Education and Sports (MoES) in Uganda (34). We chose to scale-up among S1 students because they are typically around the age of menarche (approximately 12 to 12.5 years). This optimizes benefits such as greater confidence, reduced fear and better menstrual experiences, compared with delivery to older students (5). Further, a nationwide scale-up to all classes is financially and logistically unfeasible. A multi-grade approach would duplicate content for older students, so we propose an intervention to S1 students annually.

Modelling was conducted to reflect forward looking projections under real world government delivery structures and national level needs. This model supports budgetary forecasting and planning for implementation of the MENISCUS intervention. The model utilized the ingredients based costing framework, identifying quantifying and valuing all resources required for implementation. Based on our experience in the trial, we made resource assumptions for each intervention component. For example, we assumed that four S1 teachers per school would be trained in puberty education (S1 Appendix). We included start-up and implementation costs, however, we excluded certain activities in the start-up phase to avoid duplicating existing government functions and resources (i.e. procurement of office equipment existing at MoES, exploratory school-mapping visits, procurement of branded materials (e.g. t-shirts)). We did not include staff salaries, as we assumed that the scale-up of MENISCUS activities would be integrated into routine government functions and delivered by existing staff at the MoES with no provision for the recruitment of new staff. The implementation would be guided by the National School Health Policy in collaboration with the Ministry of Health’s Division of Adolescent and School Health. The primary objective of the National School Health policy is to guide the design and the implementation of interventions to improve health in school settings in Uganda (35).

Unit costs were obtained from multiple sources, including Government of Uganda workplans and circulars, MENISCUS trial unit costs and market prices of goods and services. Required resources were estimated based on frequency and target population coverage. Costs for the potential national scale up were calculated in Ugandan Shillings (UGX) and converted to 2025 US Dollars (US$) using the mid-year exchange rate at the end of June 2025 (36).

### Sensitivity Analysis

A univariate sensitivity analyses for the national scale up were conducted on two key parameters 1) level of staff required for certain activities e.g. number of teachers per school trained in puberty education was increased from 4 to 8 teachers and 2) the number of training days for puberty education, ToTs and drama skit was increased from 2 to 4 days. Sensitivity analyses for the MENISCUS trial were reported previously (16).

### Ethics approval

We obtained ethics approval from the Uganda Virus Research Institute (UVRI) & Ethics Committee (reference GC/127/819), the Uganda National Council of Science and Technology (reference HS1525ES), and the London School of Hygiene & Tropical Medicine (reference 22952). An independent Trial Steering Committee provided scientific guidance and monitored the progress of the trial. The Independent Data Monitoring and Ethics Committee (IDMEC) reviewed the trial recruitment and safety data and provided scientific guidance. The trial was prospectively registered (ISRCTN45461276).

## Results

### Cost of a potential national scale-up of the MENISCUS intervention

When modeling potential national scale-up scenarios for all 2,995 secondary schools in 146 districts, the estimated incremental start-up cost is estimated at US$414,239 in both the basic and enhanced scale-up scenarios, and the estimated incremental implementation costs are US$9,810,446 and US$16,134,884 respectively. This results in total costs of US$10,224,685 for the basic scenario and US$16,549,123 for the enhanced scenario (Table 2). The highest cost drivers are the MH kit and training (especially in the enhanced scale-up), pain management, MH Action group (enhanced scale-up only) (Table 2).

**Table 2:** Total cost of setting up and implementing the basic and enhanced scale up scenarios.

| Activities | Basic scale-up<br>(USD) | Enhanced scale-up<br>(USD) |
| --- | --- | --- |
| ***Start-Up Cost*** |  |  |
| Planning Workshop | 7,926 | 7,926 |
| Stakeholder engagement workshops | 296,667 | 296,667 |
| Printing of the ToTs handbook and the puberty education handbook | 103,991 | 103,991 |
| Training of national level trainers | 5,656 | 5,656 |
| ***Sub-Total*** | **414,239** | **414,239** |
| ***Implementation Cost*** |  |  |
| Puberty Education | 1,521,157 | 1,521,157 |
| MH Action Group | N/A | 2,980,291 |
| Menstrual health kit and training | 2,979,874 | 6,084,064 |
| Pain Management | 2,075,559 | 2,212,382 |
| WASH improvements | 1,710,240 | 1,710,240 |
| Drama skit | 1,523,616 | 1,626,750 |
| **Sub-Total** | **9,810,446** | **16,134,884** |
| **TOTAL** | **10,224,685** | **16,549,123** |

Considering the costs of implementation only, the unit cost of scaling-up the MENISCUS intervention is estimated at US$58 per female S1 student and US$28 per S1 student (male and female) in the basic scale-up scenario and US$95 per female S1 student and US$46 per S1 student in the enhanced scale-up scenario. The unit cost per school is US$3,320 in the basic and US$5,460 in the enhanced scale-up scenario respectively.

When we consider both start-up and implementation costs, the unit cost is US$60 per female S1 student and US$29 per S1 student in the basic scale-up scenario and US$97 and US$48 in the enhanced scale-up scenario. The unit cost per school is U$3,460 in the basic, and US$5,600 in the enhanced scale-up scenarios respectively (Table 3).

**Table 3:** Total Cost per beneficiary.

|  |  | Basic scale Up |  | Enhanced scale up |  |
| --- | --- | --- | --- | --- | --- |
| Unit | Output<br>units | Unit Cost<br>(UGX) | Unit Cost<br>(USD) | Unit Cost<br>(UGX) | Unit<br>Cost<br>(USD) |
| Total cost per Senior 1 student<br>(implementation cost only) | 347,529 | 101,790 | 28 | 167,410 | 46 |
| Total cost per female Senior 1<br>student (implementation cost only) | 170,595 | 207,362 | 58 | 341,041 | 95 |
| Total incremental cost per school<br>(implementation cost only) | 2,955 | 11,971,201 | 3,320 | 19,688,599 | 5,460 |
| Total cost per Senior 1 student<br>(start up and implementation cost) | 347,529 | 106,088 | 29 | 171,708 | 48 |
| Total cost per female Senior 1<br>student (start up and<br>implementation cost) | 170,595 | 216,118 | 60 | 349,796 | 97 |
| Total incremental cost per school<br>(start up and implementation cost) | 2,955 | 12,476,676 | 3,460 | 20,194,075 | 5,600 |

### Sensitivity analysis

For the national scale-up sensitivity analysis, doubling the number of teachers participating in puberty education training per school from 4 to 8 resulted in increased incremental implementation cost in the basic (15%) and enhanced scale-up (9%) scenarios respectively. Increasing the number of training days for puberty education, TOTs and drama skit increases the incremental implementation cost substantially by 37% and 23% in the basic and enhanced scale-up scenarios respectively (S4 Table).

## Discussion

To our knowledge, this is the first paper to estimate the costs of nationally scaling-up a MH intervention to improve the health and wellbeing of in-school adolescents. Our findings offer context-specific data to inform policy decision making, budget planning and prioritization of menstrual health within the broader health and education agenda at national level.

This study presents the costs of scaling-up the intervention to all secondary schools in Uganda. We found that the MH kit and training, pain management and puberty education constituted the highest cost in the basic scale-up scenario whereas MH kit and training, menstrual health action group and pain management registered the highest cost in the enhanced scenario. There was a 64% increase in implementation costs in the enhanced scenario compared to the basic scale-up scenario.

When scaled-up nationally, the intervention benefits from economies of scale (18,37), with unit costs per school in both the basic and enhanced scenario and unit costs per student in the basic scale up scenario declining as delivery is expanded across all secondary schools in the country. This pattern suggests that efficiencies may be gained through large scale implementation and integration of existing education programs (38–40). The enhanced scenario shows mixed results of the economies of scale. When we consider implementation costs only, the enhanced scale-up scenario showed some diseconomies of scale as the unit costs of both S1 students and female S1 student increased by US$2 and US$10 respectively in comparison to unit costs per participant in the setting of the MENISCUS trial, however, when we consider both start up and implementation costs, we see economies of scale as the unit cost per S1 student and female S1 student decreased by US$6 and US$7 respectively. The scale-up scenarios presented are through a government-led model which would require absorption of the intervention components under the routine programming activities of the Government of Uganda. The additional costs of salaried staff in the MENISCUS trial would be eliminated since the role of implementation of the different MENISCUS components would be transferred to the existing staff employed by the Ministry of Education and Sports. During the MENISCUS trial, the intervention was implemented by WoMena, whose staff are paid higher salaries than staff who work in the Government of Uganda who would implement the MENISCUS intervention at national scale. The shift from NGO-led to Government-led delivery will require a robust quality assurance mechanism to ensure MH education retains its value when delivered by teachers. The Government of Uganda currently provides a capitation grant that is allocated per secondary school student under the Universal Secondary Education Programme (41). The grant is budgeted at US$49 per secondary school student and covers instructional material, co-curricular activities, school management, administration costs and contingency costs. In comparison, the additional unit cost per student required to implement the MENISCUS intervention at national scale represents slightly over half the existing cost per student under the basic scenario and approximately twice the cost per student in the enhanced scenario. This comparison underscores the additional resource requirements associated with the implementation of the MENISCUS intervention and highlights the need for budgetary adjustments to support integration of the intervention within existing school-based programmes.

The costs of implementing a different MH intervention in Kenya were lower compared to the MENISCUS trial. The annual cost of providing MH kits (cup or disposable pads and soap) and puberty training was estimated at US$34 per direct recipient compared to US$85 per S2 female student the MENISCUS study (20). The higher costs observed in the MENISCUS study are attributed to the larger scope and multicomponent nature of the intervention when compared to the study in Kenya which mainly focuses on provision of menstrual cups, menstrual pads, soap and puberty education to direct recipients. The MENISCUS study incorporated additional components such as the provision of analgesics and training on pain management techniques, WASH improvements, implementation of a drama skit and establishment of a menstrual health action group. These components were designed to address not only access to menstrual health products, but psychosocial well-being, reduction of stigma and creation of a supportive physical environment thereby making the intervention comprehensive and greater in intensity (16).

Sustaining the MENISCUS intervention under either scale-up scenario will require financial commitment from school level and central government level actors to ensure smooth implementation. Although the affordability of scaling up MENISCUS remains uncertain, demographic and socioeconomic trends in Uganda and other LMICs reinforce the importance of timely, sustained and coordinated co-investment in adolescent health and wellbeing. Evidence increasingly suggests that interventions addressing multiple dimensions of wellbeing can produce benefits that extend beyond immediate health outcomes, including improved future adult health and future economic gains. These considerations highlight the need for stronger economic evidence to guide policy decisions on the prioritization, design, and co-financing of adolescent health programmes (42).

The bulk of the financial commitment will likely have to come from the Government of Uganda central budget allocation to the different relevant ministries/departments that will support the implementation of the intervention such as the MoES, the Ministry of Health, Ministry of Water and Environment and the Ministry of Gender Labor and Social Development rather than through donor financing given the irregular flows and significant reduction of external funding to African countries (43,44). While donor funding could play an important role in the start up phases, long-term sustainability will depend on predictable domestic resource mobilization and integration of MENISCUS activities into routine sectoral budgets.

The allocation of funds to the different sector ensures that the multisectoral value of the intervention is captured and shared goals achieved efficiently (45). Certain activities such as the implementation of the drama skit performance in the enhanced scenario will have to be school-led and implemented by school administrations. The government will provide broad supervision of school level activities through existing decentralized service delivery i.e. the district structures of administration.

For policy makers to adopt and implement a MH intervention, it must be affordable and the benefits to the targeted population must be well laid out. Reliance on single-sector health budgets may be insufficient to support scale-up, this underlines the relevance of co-financing approaches that link investments across education, health, and social protection systems. Although this study did not include a formal budget impact analysis, the micro-costing approach to scale up provides detailed and context-specific estimates of the resources required for each programme component for a national scale up under different possible scenarios. By disaggregating costs at the activity level, this method identifies key cost drivers and offers insights into where efficiencies may be achieved through programme adaptation or integration within existing service delivery systems. This level of detail supports informed assessment of the feasibility of shared financing arrangements and strengthens the evidence base needed to promote the sustainability and scalability of adolescent health interventions (46–48). MH services are often not prioritized in health investments despite showing evidence of the substantial benefits to girls and women in low resource settings (8,10,12,49,52–54). The study addresses a critical gap and contributes to the growing investment case for integrating menstrual health services within national systems.

### Limitations

As with many trials that evaluate interventions in controlled settings, the effectiveness of the intervention also remains uncertain when scaled up. There is evidence to show that intervention effects tend to diminish when scaled up beyond trial settings (55). This may arise due to limited resources, reduced implementation fidelity, complexities of the intervention, change in delivery mechanisms and lack of engagement of local implementors. In the scenarios presented, we do not model the effectiveness of the intervention when scaled up nationally. In addition, the effectiveness of the MENISCUS intervention under pared back delivery models such as the basic scale up scenario that was presented here is unknown as we excluded certain intervention components that were part of the trial. Finally, we used a provider perspective (consistent with economic evaluation guidelines) and did not consider broader societal costs such as the opportunity costs for students and out-of-pocket expenditures for students and households, which may underestimate the total costs of implementing the MENISCUS intervention. A societal costing perspective may best estimate comprehensive costs including those for the user for instance out of pocket expenditure on supplementary products such as soap, pain relief items beyond those provided in the kit as well as the ongoing maintenance costs on reusable items such as water and soap for washing reusable pads Estimating these costs for this study would be challenging as out-of-pocket costs on MH are sometime small, frequent, non-monetary or paid in kind with no receipts and would require students to recall these expenses accurately, leading to recall bias and unreliable estimates (56). To conduct the scale-up, we faced challenges obtaining the most recent information on the number of secondary schools and number of students in S1 from official sources. At the time of writing this paper, a nationwide baseline education census is ongoing with no timeline provided for the results of the census. Instead, we used data from online sources from the MoES (34).

### Conclusion

In the multicomponent MH intervention trial, the main cost drivers were the salaried staff and supplies (16). For the national scale up, the main cost drivers were supplies (per-diems for national level officials, venue hire, transport refund and fuel). Despite the projected substantial costs for the startup and implementation phases during the national scale-up, interventions are essential to improve MH care for in-school female adolescents. The study provides a blueprint of cost estimates for countries intending to adopt and implement a menstrual health intervention that addresses physical and psychosocial aspects of menstrual health. The modelling of two scale-up scenarios provides policy makers with different implementation pathways for aiding decision making and financial planning with limited resources. The initial costs (startup costs) are smaller than the implementation costs, indicating the need for substantial annual implementation costs to implement the MENISCUS intervention. Using existing government structures will significantly reduce the cost of implementation by eliminating costs brought about by running parallel systems. To foster efficiencies and ensure minimization of costs, governments should consider integrated systems of delivery.

## Data Availability

The data will be available on the LSHTM repositary

## Acknowledgments

We extend our thanks to the staff, students, and communities of participating schools for their engagement with the study; the stakeholders from the Uganda Ministry of Education and Sports, District Education Officers in Wakiso and Kalungu districts, the MENISCUS research team and WoMena Uganda.

## Author Contribution

Conceptualization, funding acquisition and designed methodology: Helen A Weiss, Giulia Greco and Fred Matovu Data Collection: Stephen Lagony, Daria Bucci, Rebecca A.K.Dwommoh and Edward Obicho Data analysis: Stephen Lagony, Daria Bucci, Rebecca A.K.Dwommoh, Levicatus Mugenyi, Kate A Nelson, Giulia Greco and Helen A Weiss Project administration: Stephen Lagony, Shamirah Nakalema, Giulia Greco and Helen A Weiss Writing: Stephen Lagony, Daria Bucci, Rebecca A.K.Dwommoh, Levicatus Mugenyi, Kate A Nelson, Edward Obicho, Fred Matovu, Shamirah Nakalema, Helen A. Weiss and Giulia Greco.

## S1 Appendix Scale up of the MENISCUS Intervention

### I) Startup activities

The following will be conducted as part of startup activities;

#### 1. Planning meeting

A national planning workshop will be convened to develop a road map for the scale up of the MENISCUS intervention to all the 146 districts in Uganda. The meeting will be organized by Ministry of Education officials and will involve approximately 60 officials from all relevant ministries such as the Ministry of Water and Environment, Ministry of Gender, Labor and Social Development, Ministry of Local Government, Ministry of Health, Ministry of Works and Transportation and the Ministry of Finance, Planning and Economic Development. We anticipate that the workshop will be held over two working days and will require hiring a venue, meals & refreshments, stationery, printing and a transport refund for all participants.

#### 2. Stakeholder engagement workshops

This activity involves sensitization and planning with all relevant stakeholders from all districts to conduct large-scale implementation of the MENISCUS intervention. Using the current Government of Uganda cascaded model of national training and engagement, we anticipate that there will be 17 regional level stakeholder engagement workshops in the different sub regions of Uganda namely; Acholi, Ankole, Bukedi, Buganda Bugisu, Bunyoro, Busoga, Kampala, Karamoja, Kigezi, Lango, Madi, Rwenzori, Sebei, Teso, Tooro, and West Nile. District officials within each of these regions will convene in a central district location of each region to be sensitized about the wide scale implementation of the intervention. Various district level officials such as the Chief Administrative Officers (CAOs), the district chairperson LC V, Resident District officials (RDCs), District Education Officers (DEOs), District Health Officers (DHOs) and the District Inspector of schools will be invited for a one-day stakeholder engagement. School level officials such as the head teacher, director of studies, and health facility in charges will also be invited to the regional meetings. The 17 different meetings will require: hiring a venue, meals & refreshments, stationery, printing, fuel for national level officials, lodging for participants, and transport refund for all district participants.

#### 3. Training of national level trainers

The training of national level trainers will be centralized and conducted in Kampala district and thereafter 3 national level trainers will be distributed to each of the regions to conduct regional level ToTs, Menstrual Health (MH) action group trainings and puberty education. A total of 51 trainers and 10 MoES officials will be trained to adequately cover all the 17 regions. The ingredients required for this training include meals and refreshments for the participants, hiring a venue, transport refund, a trainers fee and stationery.

#### 4. Printing of the training of trainer handbook and the puberty handbook

The training of trainer handbook was adopted to the MENISCUS intervention before the trial was conducted and for the scale up, we assume the use of the same handbook to conduct the Training of Trainers (ToT) and Training of Beneficiaries (ToB). For the 30 schools, 230 handbooks were printed to facilitate the Training of trainers (ToT) and the Training of Beneficiaries (ToB) in Wakiso and Kalungu districts. In all the 2,995 secondary schools, we estimate that a total of 11,481 handbooks will be needed for all the training sessions. We shall cost an extra 10% of the required books, to account for possible losses and destruction in some cases.

The Government of Uganda puberty handbook will also be printed to facilitate the puberty education training sessions to the teachers. One handbook per school will be printed and distributed to the schools during the regional puberty education training sessions to teachers.

### II) Implementation Activities

The following activities will be conducted as part of the implementation of the scale up of the MENISCUS intervention to national level. For each of the activities, we differentiate the activities that fall under the different scenarios;

#### 1. Puberty education

In this activity, schoolteachers will be trained on how to deliver the Ministry of Education and Sports’ (MoES) training on menstrual health management. After the training of national level trainers in the start-up phase, the 51 trainers will be distributed amongst 17 sub-regions (3 in each region) to conduct puberty education training to schoolteachers from the schools in every district. The puberty education training sessions will be conducted over two days and will include an introduction to the intervention, discussion of the puberty education curriculum, and sessions on delivering government MHM school guidelines. The trained teachers will then routinely conduct puberty education sessions to both male and female students in their classrooms. We assume that an average of 4 S1 teachers will be trained from every school who will thereafter deliver puberty education sessions as part of their routine teaching activities in their classrooms. The key ingredients required for the puberty training include hiring of 17 training venues in the different sub-regions of Uganda, meals and refreshments for the participants, lodging for the teachers and trainers, stationery, a transport refund for the teachers and fuel for district and government officials. These inputs will be similar across both scenarios.

#### 2. Menstrual Health (MH) Action group

In the enhanced scale up scenario, a two-day training for selected MH Action group members from the different secondary schools will be conducted. Teachers and a select group of students from each school will be included to serve as trainers during the training of beneficiaries before the distribution of the reusable pads and underwear and simultaneously as members of the MH Action groups. A total of 8 members per school will be selected to take part in the MH action group training. The people who would’ve been trained during the MHM action group trainings at the regional level trainings will be expected to integrate menstrual health activities as part of their routine club activities. The key ingredients required for this activity include hiring 17 training venues for each of the regions, procurement of a demonstration kit to be used in the training sessions, meals & refreshments, stationery, transport refund for participants, lodging for participants and fuel for central and district level officials.

#### 3. MH kit & training

For the basic and enhanced scale up scenarios, a two-day training session (ToTs) will be conducted for school-level trainers to equip them with the skills needed to deliver training of beneficiary sessions to S1 female students within their schools. Teachers and a select group of students from each school will be included to serve as trainers during the training of beneficiaries before the distribution of the reusable pads and underwear and simultaneously as members of the MH Action groups. In the enhanced scenario, the menstrual health kit which contains re-usable Ugandan-manufactured menstrual pads (AFRI pads) and underwear will be procured and distributed to female S1 students. These items could be procured through the Uganda National Medical Stores that has the mandate to procure, store and distribute human medication and health-related consumable items to all government owned health facilities in the country. These items can then be accessed by the schools from nearby health facilities and later distributed to the female students in the schools. Lastly, for the enhanced scale up scenario, a one-day training of female care givers will be conducted with 8 female care givers from each of the schools in the 17 sub-regions of Uganda. The requirements for the training sessions include, meals & refreshments, hiring a venue for the training, lodging for the participants, fuel for central and district level government officials, stationery and a transport refund.

#### 4. Pain Management

For both the basic and enhanced scale up scenarios, this activity will include training on effective pain management strategies to manage period pain. The training will be delivered to select school-based service providers such as senior women and men, nurses, teachers etc. This training will be conducted over the course of 1 day. In addition, the enhanced scale up scenario will include provision of 6 tablets of ibuprofen per month to female students in S1.

#### 5. Water, Sanitation and Hygiene (WASH) improvements

WASH improvements as part of the trial involved fixing broken toilet doors and locks, provision of sanitary bins, toilet paper cages, water carriers/stand, provision of liquid soap (5L), and (20L) water containers. For the basic and enhanced scale up scenarios, it’ll involve hiring of a contractor who will carry out the WASH improvements in all secondary schools in Uganda such as procurement of WASH stands, drums and basins, fixing broken toilet doors, installing toilet paper cages and door locks.

#### 6. Drama skit

During the implementation of the MENISCUS trial, the school drama club was assisted by WoMena to implement the drama skit. WoMena trained the school drama club members on the implementation of the skit before a performance was conducted for all students in the school. The implementation of the drama skit is a school level activity and it will involve the adaptation of the already developed drama script by the school drama clubs. For both the basic scale up and the enhanced scale up scenarios, the drama script will be printed and distributed to each school. Drama skit rehearsals will be conducted through training sessions in all the 17 regions. School representatives from drama clubs from all the schools in Uganda will be invited to regional drama skit rehearsal sessions. The rehearsal sessions will be conducted over two days by trainers from the Ministry of Education and Sports. These will be done in the form of training workshops where school representatives will familiarize themselves with the drama script under the direction of the trainers from the Ministry of Education and Sports. The school representatives will then lead the drama club members in their respective schools in practice and school club rehearsals. The regional sessions will also include district representatives such as the district inspector of schools and district educator officers from all the districts. In the enhanced scenario during the quarterly inspection of schools by the district inspector of schools, checks will be done to determine whether the drama skit performances are regularly conducted. S1 Table shows the resources and assumptions used.

**S1 Table:** Assumptions and resources for start-up and implementation activities for the scale up of the MENISCUS intervention.

| Start-up phase | Resources | Assumptions | Sources of costing information |
| --- | --- | --- | --- |
| Planning meeting | Venue, meals & refreshments, | 60 officials from Ministry of Education and Sports, Ministry of Gender, Labor and Social | MENISCUS trial costs, Government of Uganda activity work plans and the |
|  | transport refund, stationery, printing | Development, Ministry of Finance, Ministry of Water, Office of the Prime Minister, Ministry of Local Government and the Ministry of Health and other relevant government bodies | duty facilitating allowances for public officers |
| Stakeholder engagement workshops | Venue, meals & refreshments, fuel, transport refund, lodging for participants, printing stationery | A one-day meeting will be conducted in the 17 sub regions of Uganda with 20 participants from each of the 146 districts attending. 100 liters of fuel will be provided to 3 Ministry of Education officials to travel to each region to conduct the stakeholder engagements. | MENISCUS trial costs, Government of Uganda activity work plans and the duty facilitating allowances for public officers |
| Training of national level trainers | Meals & refreshments, venue, stationery, trainer fee for trainers and transport refund | One training meeting to be conducted in a central location. A total of 51 trainers and 10 MoES officials will be trained for 2 days. Every region will receive three trainers to conduct a | MENISCUS trial costs and the Government of Uganda duty facilitating allowances for public officers |
| Printing of the Training of Trainer handbook and the puberty education handbook | Training of trainer Handbooks<br>Puberty education handbook | Number of handbooks were determined by the number of handbooks printed for the 60 schools during the MENISCUS trial. We costed an additional 10% of the required number to account for possible losses and destruction | MENISCUS trial costs |
| **Implementation phase** | **Resources** | **Assumptions** | **Sources of costing information** |
| Puberty education | Venue for puberty education trainings in 17 regions, meals & refreshments, lodging for participants, stationery, transport refund, fuel, and stationery | Puberty education trainings will be conducted in 17 regions. 4 senior one teachers will be trained from each of the schools over 2 days. | MENISCUS trial costs, Government of Uganda activity workplans, prevailing market prices for fuel and duty facilitating allowances for public officers |
| MH Action Group | Venue, demonstration kits to be used in the training sessions, meals and refreshments, printing, stationery, lodging for participants, fuel and transport refund | 8 representatives per school shall be trained at regional level. 3 trainers will be involved in conducting MH Action group training. 3 district officials per district will also be involved in the trainings. | MENISCUS trial costs, Government of Uganda duty facilitating allowances for public officers, prevailing market prices for fuel and activity workplans |
| Menstrual Health Kit and training | Venue hire, meals & refreshments for participants, lodging, stationery, transport refund and fuel for central and district level government officials. | 8 selected members per school will be trained in the TOTs over 2 days, 8 selected female care givers per school will also be trained over a 1-day training. In the enhanced scale up, underwear and reusable pads will be distributed to all S1 female students | MENISCUS trial costs, Government of Uganda duty facilitating allowances for public officers and prevailing market prices for fuel |
| Pain Management | Venue to conduct pain management training, meals & refreshments, lodging for | 8 participant per school will be trained on pain management techniques. An option of ibuprofen will be procured for | MENISCUS trial costs, market prices, Government of Uganda activity workplans |
|  | participants, stationery, transport refund and fuel. Ibuprofen and drug envelopes will also be provided to female S1 students in the enhanced scale up scenario | all female students who may require pain killers to manage pain during their periods. |  |
| WASH Improvements | Procurement of WASH stands, drums and basins, fixing broken toilet doors, installing toilet paper cages and door locks | All schools will require WASH improvements | MENISCUS trial costs |
| Drama skit | Printing of drama script, Venue, meals & refreshments, lodging for participants, stationery, transport refund and fuel | 4 drama club members per school will be trained at regional level trainings | MENISCUS trial costs |

##### Total cost of setting up and implementing the MENISCUS Intervention in the trial

The total economic cost of the MENISCUS intervention in 30 schools of Wakiso and Kalungu districts during the MENSICUS trial was US$222,493. This comprised the set-up cost of US$40,990, and the annual implementation cost of US$181,503 (S2 Table). Salaried staff accounted for approximately 43%, and supplies for 34% of the total cost (16).

**S2 Table:** The Cost of Setting Up and Implementing the MENISCUS Intervention in the trial.

| | Resources used | US\$ | % of<br>total cost |
| --- | --- | --- | --- |
| **Start Up Phase** | Equipment | 1,114 | 0.50 |
|  | Buildings: Spaces | 2,861 | 1.29 |
|  | Buildings: Furniture | 66 | 0.03 |
|  | Salaried Staff | 31,384 | 14.11 |
|  | Supplies | 2,573 | 1.16 |
|  | Building Utilities & Maintenance | 1,055 | 0.47 |
|  | Transport: Public Transportation/Rental | 1,316 | 0.59 |
|  | Per Diems and Allowances | 621 | 0.28 |
|  | **Subtotal:** | **40,990** | **18.42** |
| **Implementation**<br>**Phase** | Equipment | 4,641 | 2.09 |
|  | Buildings: Spaces | 5,740 | 2.58 |
|  | Buildings: Furniture | 783 | 0.35 |
|  | Salaried Staff | 64,922 | 29.18 |
|  | Supplies | 73,337 | 32.96 |
|  | Building Utilities & Maintenance | 1,645 | 0.74 |
|  | Transport: Vehicle Operations (fuel) | 2,763 | 1.24 |
|  | Transport: Public Transportation/Rental | 16,859 | 7.58 |
|  | Per Diems and Allowances | 10,813 | 4.86 |
|  | **Subtotal:** | **181,503** | **81.58** |
|  | **Total:** | **222,493** | **100.00** |

Reproduced from: Nelson KA, Lagony S, Kansiime C, Torondel B, Tanton C, Ndekezi D, et al. Effects and costs of a multi-component menstrual health intervention (MENISCUS) on mental health problems, educational performance, and menstrual health in Ugandan secondary schools: an open-label, school-based, cluster-randomised controlled trial. The Lancet Global Health. 2025 May 1;13(5):e888–99. doi: 10.1016/S2214-109X(25)00007-5.

##### Unit Cost of the MENISCUS Intervention

When only implementation costs are considered, the unit cost per S2 student was US$44, and the cost per female S2 student was US$85. The unit cost of implementing the intervention was US$6,050 per school. When we consider both start-up and implementation costs, the unit cost was US$54 per S2 student, US$104 per female S2 student with the unit cost per school at US$7,416 (16).

##### Implementation cost per intervention component

When examining the implementation cost per intervention component, the MH kit accounted for 36.2% of the total cost, followed by the MH action groups (16.2%), drama skits (11.0%), WASH (11.0%), puberty education (6.3%) and pain relief (3.5%). Management and administration costs accounted for 12.5% of the total implementation costs (S3 Table).

**S3 Table:** Implementation cost per intervention component (USD).

| Intervention component | Implementation cost (USD) | % of total |
| --- | --- | --- |
| Cross-cutting activities | 4,759 | 2.6% |
| Puberty education | 11,383 | 6.3% |
| Drama skit | 19,939 | 11.0% |
| Menstrual health kit | 65,745 | 36.2% |
| Pain relief | 6,416 | 3.5% |
| Water, sanitation, and hygiene facilities | 21,047 | 11.6% |
| Action groups | 29,484 | 16.2% |
| *Management and administration* | *22,729* | *12.52%* |

Reproduced from: Nelson KA, Lagony S, Kansiime C, Torondel B, Tanton C, Ndekezi D, et al. Effects and costs of a multi-component menstrual health intervention (MENISCUS) on mental health problems, educational performance, and menstrual health in Ugandan secondary schools: an open-label, school-based, cluster-randomised controlled trial. The Lancet Global Health. 2025 May 1;13(5):e888–99. doi: 10.1016/S2214-109X(25)00007-5.

**S4 Table:** Sensitivity Analysis results for trial and scale up.

| Parameter Varied | Base Case | Variation | % Change in Costs |
| --- | --- | --- | --- |
| Lifespan of start-up activities | 1.5 years | 3 years | –48% |
| Shadow price of menstrual cup | US\$7.25 | US\$20 | 4% |
| Discount rate | 11% | 3–7% | ±1% |
| Exchange rate | US\$/UGX 3,662 | Lowest–highest 2023 | ±2–4% |
| Teachers trained per school | 4 | 8 | +9–15% |
| Training duration | Base | +2 days | +23–37% |

